# Frequency and Risk Factors of Resident Burnout Before and During the COVID19 Pandemic

**DOI:** 10.1101/2022.09.04.22279366

**Authors:** Sevcan Türk, Anıl Kalyoncu, Özlem Sürel Karabilgin Öztürkçü, Hayriye Elbi, Mehmet Nurullah Orman, Aziz Cem Yaman, Muratcan Toptaş, Yaşar Çağlar Bekki, Murat Özlütaş, Mehmet Burak Esentürk, Jayapalli Bapuraj, Süha Süreyya Özbek

## Abstract

**Objective:** To explore the prevalence and contributing factors of resident burnout in a University Hospital before and during the COVID 19 pandemic.

**Methods:** Thirty Faculty of Medicine departments were included in the survey, where 400 university hospital residents filled out the Maslach Burnout Inventory (MBI) in January 2018 and April 2020. Related scores of emotional exhaustion (EE), decreased accomplishment (DA) and depersonalization (DP) were calculated and compared between the different groups. Correlation between scores and possible contributing factors, including demographics, work-life circumstances, exposure to workplace violence, were investigated. Contributing factors were compared between the time points.

**Results:** The EE and DA scores were significantly higher in junior residents than in senior residents. Both scores were higher among residents who had experienced abuse or violence. The emergency medicine residents had significantly higher DP scores, while the EE scores of radiology residents were lower than others. Thirty percent of all residents smoked cigarettes. This percentage was even higher among the residents of the departments of surgery and emergency medicine (45-50%). A significant correlation was demonstrated between the scores of MBI and smoking, while analysis with other demographics did not yield any relation. According to the study results, basic science residents had significantly increased scores in all MBI subgroups during the pandemic.

**Conclusion:** Resident physician burnout was found to be related to the work environment, smoking cigarettes and exposure to violence at workplace.

## Background/Introduction

Maslach has described burnout as a condition in which professionals “lose all concern, all emotional feelings for the people they work with and come to treat them in a detached or even dehumanized way” (1, 2). There are three main components of burnout. (1) feelings of cynicism or depersonalization (2) a sense of ineffectiveness, and (3) lower efficacy. Burnout in medical staff has been reported to range from 18.7% to 74.8%. The prevalence of burnout was highest in Asian countries 57% followed by North America at 51% and 27% in European countries(3). Physicians, especially those exposed to high levels of work-related stress, have a reportedly higher prevalence (4). Surveys among practitioners from family practice, internal medicine, surgery, and the emergency room, show low levels of motivation, fear of overwork, rising caseloads, and increased sbureaucracy as the leading causal events for burnout. This has resulted in a growing number of physicians leaving the medical profession, citing mental and physical health issues (5, 6). Physicians also have one of the highest suicide rates amongst professionals (5, 6). This could be explained by the high prevalence of burnout, the administrative burden with insufficient autonomy, and collegial conflicts (7). Professional fulfillment, emotional exhaustion, and interpersonal disengagement were significantly associated with burnout in a survey study suggesting the predictors of burnout among a group of radiology faculty (8). Sleep disorders, reduced self-compassion, negative work impact on personal relationships, lack of perceived appreciation for work, lack of control of the schedule, lack of personal-institutional values alignment, and reduced leadership quality were other important factors in the same study (8). Therefore, this study aimed to ascertain the prevalence of burnout among residents at our institution and evaluate its relationship with possible contributing factors before and during the Covid 19 pandemic.

## Materials and Methods

This cross-sectional survey was conducted twice in January 2018 and then in May 2020 at a University Hospital. The University institutional review board approved the study. Consent forms were attached to the survey, which was filled by participants anonymously. Formal consent to carry out this research was also obtained from each department chair as per the rules of the Faculty of Medicine.

Randomly selected two hundred respondents representing the whole group were included in the surveys at each of the two-time points of the study, with a total of 400 completed with a 100% response rate. 117 internal medicine residents, 38 surgery, 21 basic science, and 24 emergency medicine residents were included in 2018; and 113 internal medicine residents, 47 surgery, 11 basic science, and 28 emergency medicine residents were included in the 2020 survey (Table 1). The total strength of the resident population in the hospital is 600. The data of marital status, being a single parent, living alone or with family, the salary, smoking, exercising and sleeping habits, medical history, and daytime and night shift hours were included in the demographic survey. MBI questionnaires were collected separately. The questionnaire was distributed and collected by medical students assigned to the residents’ clinical rotations in 2018. The survey was filled online by the residents in the coronavirus pandemic by using Surveymonkey as the online data gathering tool. The security and confidentiality of the information contained in the survey was monitored and secured at every step of this study.

**Table 1:** Residents filling the survey are 30% (200/600) of all residents at the hospital.

| Department | Survey 2018 |  | Survey 2020 |  |
| --- | --- | --- | --- | --- |
| Medial Science | 117 (%59) | 63/54 (F/M) | 113 (%57) | 64/49 (F/M) |
| Surgery | 38 (%19) | 15/23 (F/M) | 47 (%24) | 18/29 (F/M) |
| Basic Science | 21 (%11) | 14/7 (F/M) | 11 (%6) | 9/2 (F/M) |
| Emergency Department | 24 (%12) | 10/14 (F/M) | 28 (%14) | 13/15 (F/M) |
| Total number | 200 | 102/98(F/M) | 199 | 104/95(F/M) |

Data collected in 3 dimensions with MBI: emotional exhaustion (EE) (tiredness, somatic symptoms, and decreased emotional resources); depersonalization (DP) (developing negative, cynical attitudes and impersonal feelings towards the patients); and lack of personal accomplishment (PA) (feelings of incompetence, inefficiency, and inadequacy) were analyzed(6).

Related scores of emotional exhaustion (EE), decreased accomplishment (DA) and depersonalization (DP) were calculated and compared between different resident groups and for demographic parameters. Burnout was defined by one abnormal score in one or more of the three dimensions (EE, DP, or PA). The correlations between MBI scores and demographic structures, work-life circumstances, stress, and abuse were documented.

Student t-test, one-way ANOVA, Levene’s test for equality of variances, Pearson correlation, and posthoc Bonferroni tests were used for statistical analysis for each of the three dimensions.

## Results

There is no significant difference in demographic parameters correlation with MBI scores for 2018 and 2020 time points. The mean age was 28 (±2) years in 2018 and 27 (±2) years in 2020.

Sixty percent of surgery residents and 58% of emergency department residents were male, with 33% in basic sciences (Table 1). Thirty percent of residents smoking cigarettes. The percentage of smokers was significantly higher among the residents of the departments of surgery and emergency medicine.

A significant correlation was demonstrated among the DA scores of the MBI scale and smoking (p=0.04), while analysis with other parameters did not yield any relation with demographics.

76% of all residents did not exercise regularly, whereas the prevalence of lack of exercise was significantly higher at 85% amongst basic science residents. Emergency department residents had substantially higher monthly income (p=0.00) and lower proficiency exam scores than other departments. Forty percent of residents reported workplace abuse, and 46% experienced violence from patients (Table 2). Reported workplace abuse was higher in emergency department residents at 50% in 2018 and 78% during the pandemic. There was a statistically significant correlation between high EE and DP scores and reported workplace abuse (p<0.01). The emergency medicine residents had significantly higher DP and EE scores (Figure 1, Figure 2). In contrast, the EE scores of basic science residents were lower than others before the pandemic, which significantly increased in the times of pandemic (Figure 2). Depersonalization (DP) scores increased in surgery and basic science residents during the pandemic (Figure 3). The emergency department residents had the highest DP scores, which were significantly higher than any other department at all times. EE and DP scores tended to be higher in emergency medicine and surgery residents than others at all times. Moreover, those scores were significantly higher in junior residents than in senior residents, respectively (Table 3).

**Table 2:**
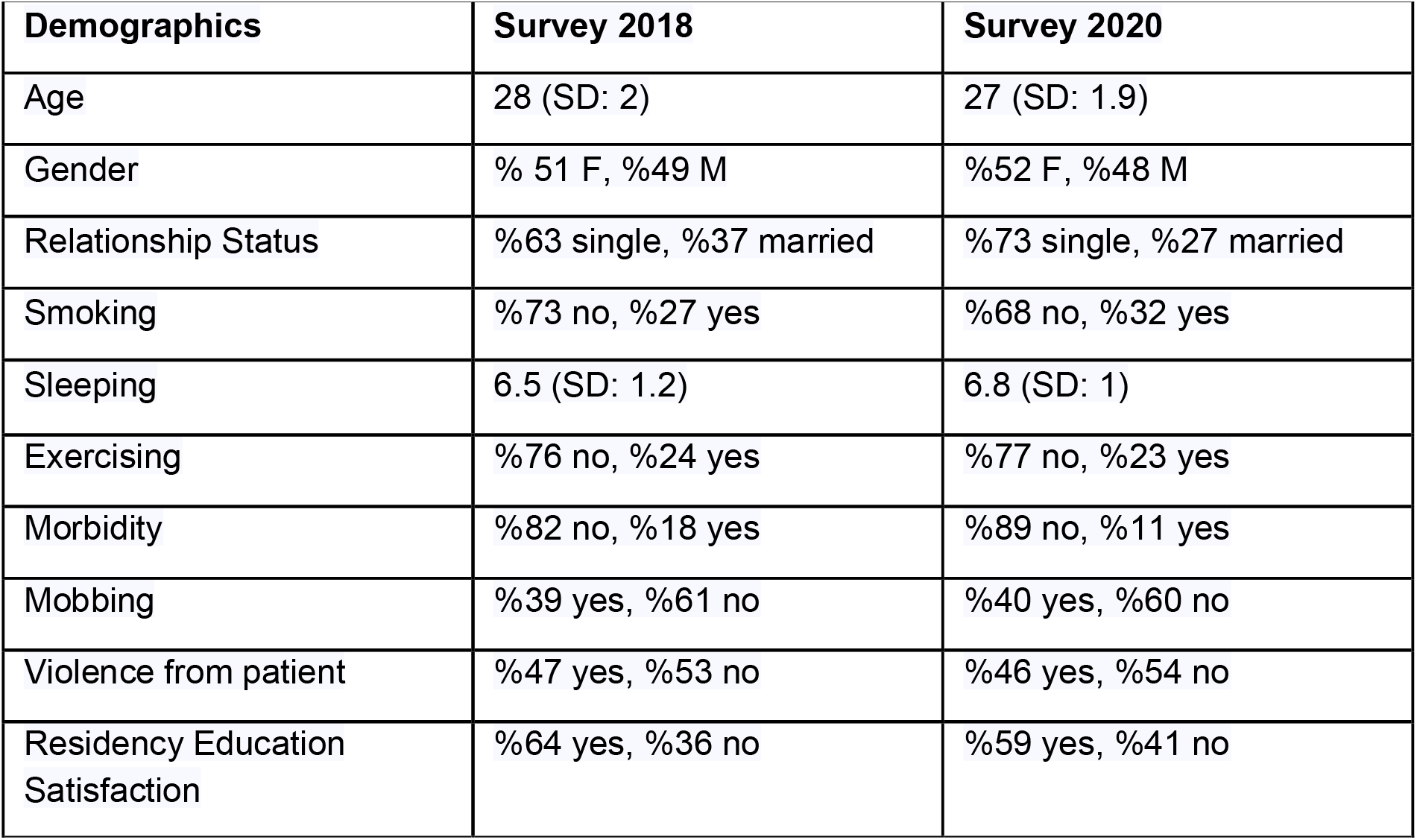
Demographic parameters among responder residents in two different time points.

**Table 3:** The EE and DA scores were significantly higher in juniors than seniors, respectively.

| Score | Age | N | Mean | Std Deviation |
| --- | --- | --- | --- | --- |
| **EE** | <=1989 | 68 | 26.5 | 7.2 |
|  | >=1990 | 93 | **28.8** | 6.6 |
| **DP** | <=1989 | 81 | 11.6 | 4.3 |
|  | >=1990 | 111 | 11.9 | 3.9 |
| **DA** | <=1989 | 76 | **29.1** | 5.5 |
|  | >=1990 | 102 | 27.1 | 4.7 |

**Figure 1:**
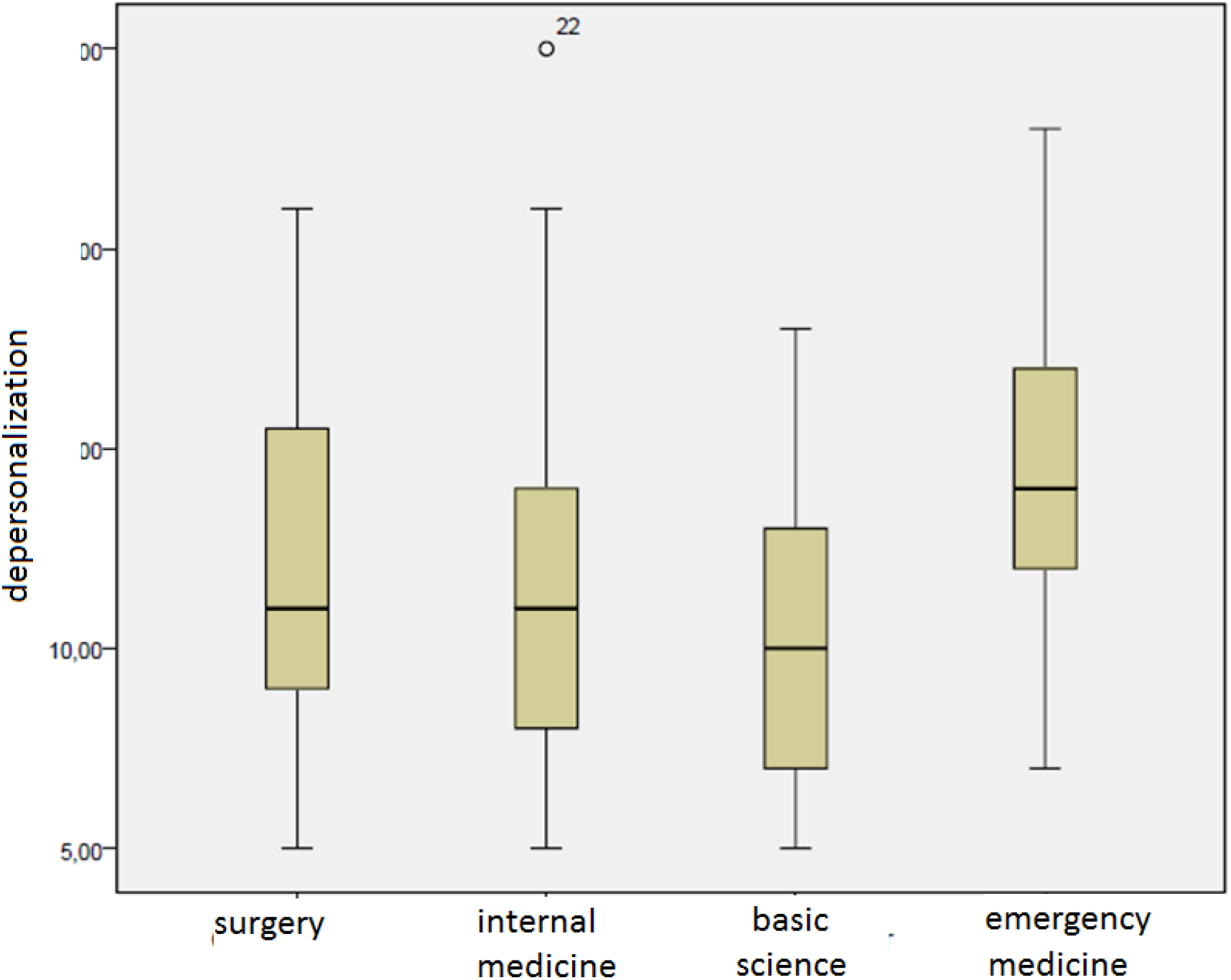
Depersonalization scores among emergency department residents were significantly higher than the others.

**Figure 2:**
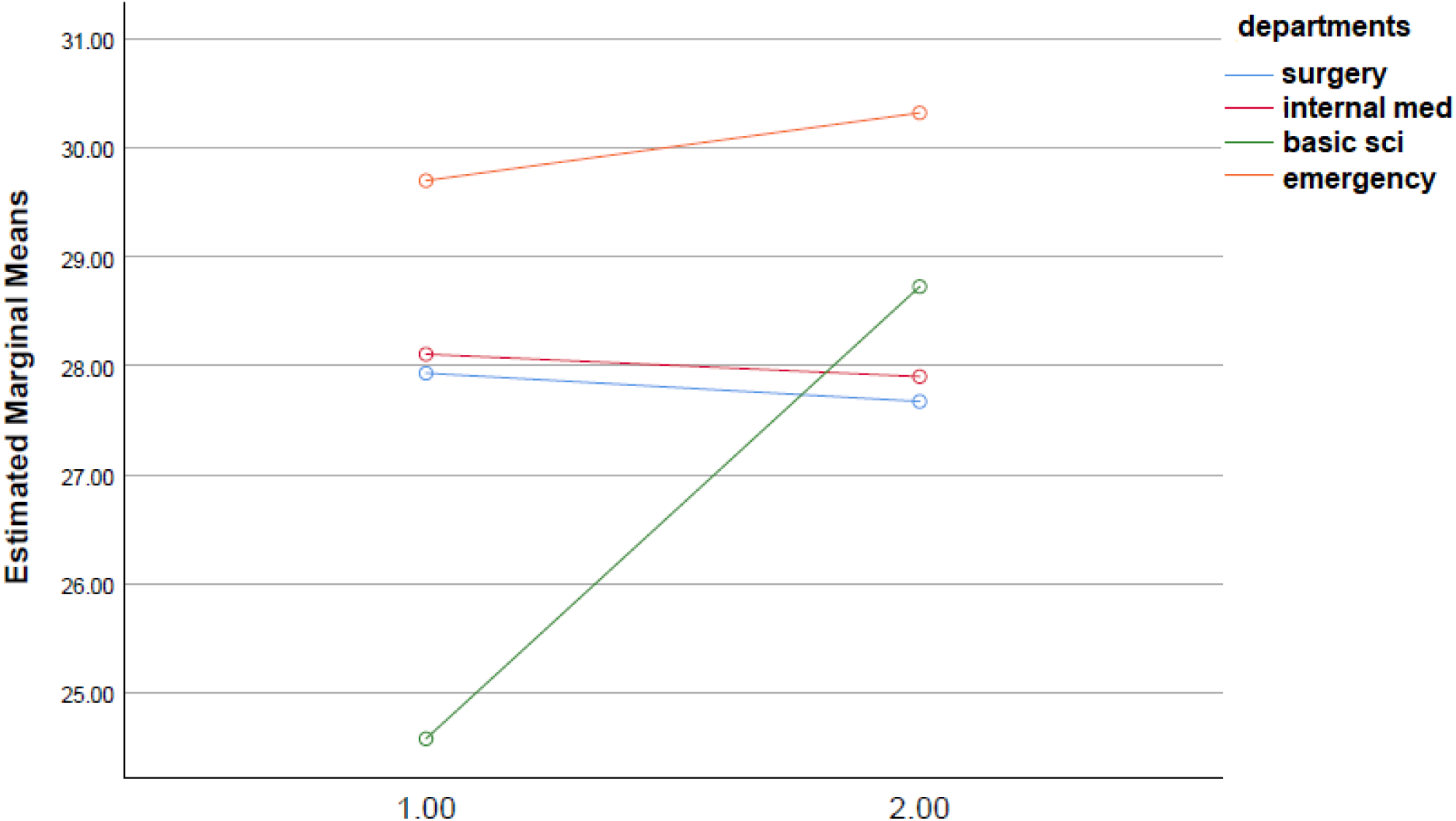
Comparison of two time points between groups for emotional exhaustion (EE). There was a statistically significant increase in basic science residents’ EE scores in the pandemic time.

**Figure 3:**
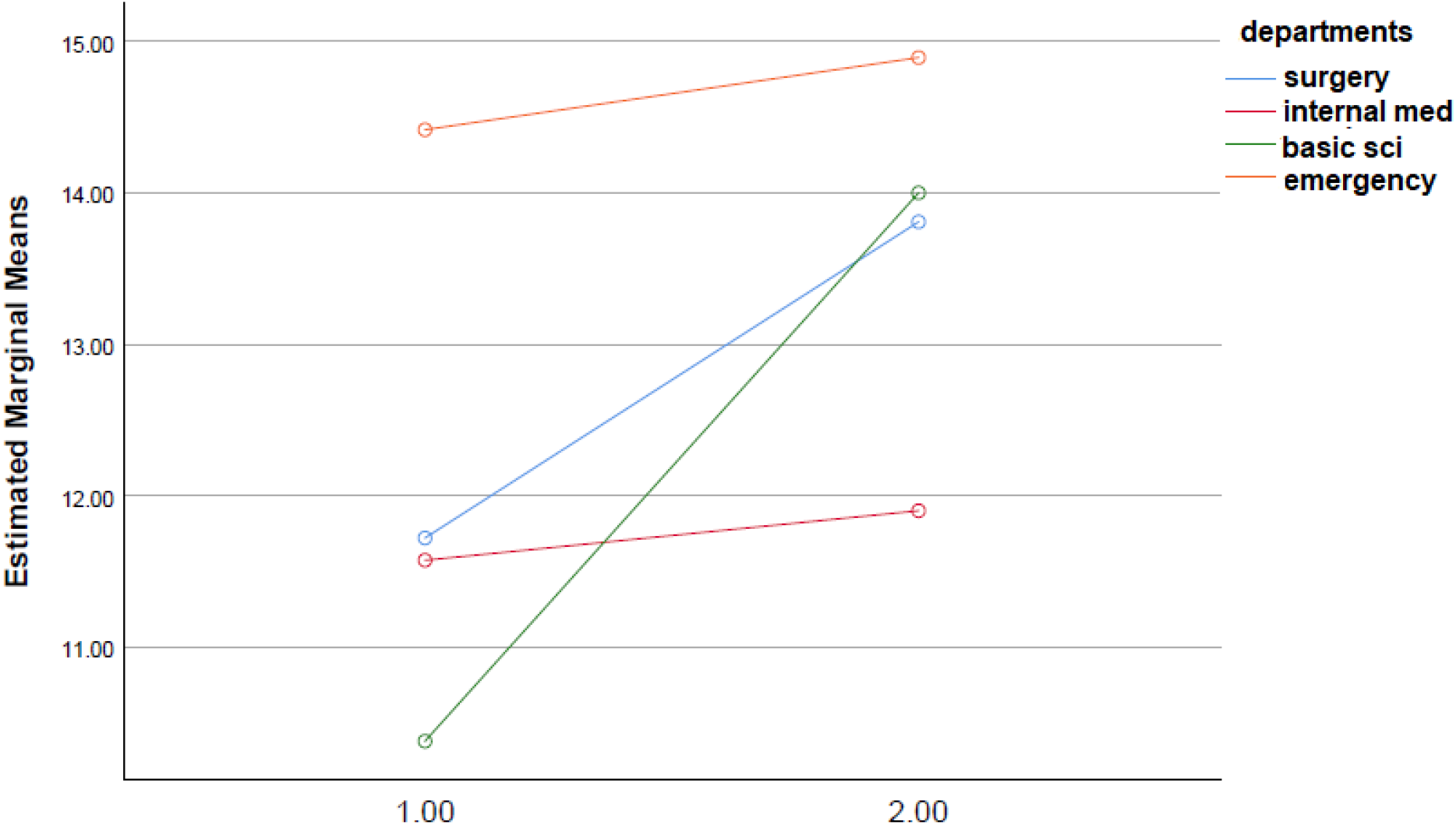
Depersonalization (DP) scores among surgery and basic science residents increased in the pandemic time. The emergency department’s residents had the top DP scores at all times which was significantly higher than any other department.

Thirty percent of residents stated that ‘they feel fatigued when they wake up every morning at the start of each working day. Twenty-five percent of residents said that ‘every day they feel working too hard on the job’. 62% (123/200) of residents were satisfied with the institution’s education, and this satisfied group of residents had significantly lower MIB scores than others (p<0.004).

## Discussion

The Covid 19 pandemic has affected health professionals in more profound ways than in other walks of life. Frontline health care professionals have faced an unprecedented situation marked by extreme mortality and morbidity. In several parts of the world, the brunt of the responsibilities of first-line medical care has been borne by resident trainees who have been recruited for the first line of encounters with patients and their immediate caregivers. Given their lack of experience and the enormity of the clinical severity of cases, work-related stress has significantly impacted the well-being of residents recruited away from their intended career paths, leading to burnout.

Burnout increasingly tends to be a concern that affects residents’ health, well-being which is significantly related to the work environment. It has been extensively associated with an increased risk of major depressive disorder, suicide, cardiovascular diseases, and substance abuse (9, 10). Leading causes of burnout include excessive workload, the imbalance between job demands and skills, a lack of job control, and prolonged work stress (11).

We evaluated resident doctors’ burnout levels by utilizing the Maslach Burnout Inventory. We compared burnout scores before and during the Covid-19 pandemic in different cohorts of students attending the same University. There was a significant difference in the two-time points in terms of emotional exhaustion and depersonalization scores for basic science residents who encountered unfamiliar working conditions, which causes a higher stress level and more exposure to workplace violence, which correlates with higher burnout scores. Basic science residents had work shifts at the emergency department during the pandemic; otherwise, all other demographics were the same between the two time points. This indicates a relationship between the working environment, workplace violence exposure, higher stress level relationship with increased burnout scores among basic science residents. Another study among emergency physicians showed a similar correlation between emotional exhaustion and exposure to violence, supporting our study’s results (12). During the Covid-19 pandemic, working hours and night shifts were novel to recently appointed basic science residents’ clinc routine, given the unfamiliar terrain of the emergency department far removed from their intended hospital work environment. Regarding work hours and night shifts, a study demonstrated that emotional exhaustion score is significantly correlated with the level of responsibility and increased occurrence of medical errors. This correlation supports that high emotional exhaustion scores among basic science residents during the pandemic can be related to increased workload and dealing with unfamiliar working conditions (13).

Concordant with the literature reviewed for this project, we demonstrated that smoking, workplace abuse, working at the emergency or surgery departments were associated with higher burnout scores. A study conducted in Australia also showed higher work stress related to increased smoking rates (14). This suggests that residents use smoking as a resilience tool, which may not be effective well. However, smoking was a good indicator of burnout among residents in our study. The higher EE scores correlated with experiencing abuse and violence, which is significantly increased in emergency medicine residents at all times. Burnout, therefore, was related to workplace environment safety.

Our study provided a framework for a closer but a widened perspective for resident wellness. The Covid 19 pandemic has opened a new pathway that may bring a sea change in the training curricula of medical resident training across specialties. One of the significant bedrock for bringing about this change is inculcating a sense of resilience and well-being amongst residents.

Resilience can be achieved from two perspectives. Physician-directed interventions typically involve mindfulness techniques or cognitive-behavioral techniques to enhance job competence and improve communication skills and personal coping strategies like educational interventions targeting physicians’ self-confidence and communication skills, exercise, or combining these features. And organization-directed interventions can involve simple changes in schedule and reductions in the intensity of workload or more ambitious changes to the operation of practices and whole health care organizations (15). Misalignment of values between the individual and the institution was significantly associated with burnout. In contrast, studies support that burnout is not an individual but an institutional and health care organization problem (16, 17). In these perspectives, organization-directed approaches that promote healthy individual-organization relationships were suggested to reduce burnout (17).

Limitations: Basic science residents were a small number of the group. However, they represented 50% percent of their department. We could not compare all departments separately because of the limited number of residents in many specialties. A randomly selected representative group of residents were reached to fill in the survey, and the response rate was 100%. The comparison was made under general groups like surgery and internal medicine, which may not represent fewer subspecialties. Conclusion

Burnout increasingly tends to be a concern that affects residents’ health, well-being, and job performance, significantly related to the work environment. This study demonstrated a relationship between a higher level of work-related stress, workplace violence, and smoking to statistically significant a higher level of EE, DP, and DA scores among residents. The DP score is significantly increased in basic science and surgery department residents in pandemic times due to the change in the work environment. Frontline health care professionals have faced an unprecedented situation marked by extreme mortality and morbidity. In several parts of the world, the brunt of the responsibilities of first-line medical care has been borne by resident trainees who have been recruited for the first line of encounters with patients and their immediate caregivers. Given their lack of experience and the enormity of the clinical severity of cases, work-related stress has significantly impacted well-being, leading to burnout.

## Data Availability

All data produced in the present study are available upon reasonable request to the authors

## Notes

Conflicts of interest: None

### Competing Interest Statement

The authors have declared no competing interest.

### Funding Statement

This study did not receive any funding

### Author Declarations

Ege University Hospital Ethics Committee approved the study. Each department gave consent for the survey before reaching the residents.

